# Pharmacokinetics of Intravaginal, Self-Administered Artesunate Vaginal Inserts Among Healthy Women in Kenya

**DOI:** 10.64898/2026.07.10.26357746

**Authors:** Annum Sadana, Sabine J Yessayan, Kashish C Patel, Martin Ongas, Brenda Misiko, Cynthia Cheserem, George Githongo, Lisa Rahangdale, Jennifer H Tang, Jerome L Belinson, Jackton Omoto, Jesse DeLuca, Daniel Selig, Kristina Pannone, Chau Vuong, Mihaela Plesa, William Zamboni, Chemtai Mungo

**Author notes:** Corresponding author: (AS).

## Abstract

**Background:** Cervical precancer (CIN2/3) remains undertreated in low- and middle-income countries due to limited access to screening and healthcare providers, contributing towards high cervical cancer mortality rates. Self-administered intravaginal therapies, such as artesunate, may help to expand access to cervical precancer treatment. However, data on the systemic absorption and pharmacokinetics of intravaginal artesunate are lacking.

**Objective:** This study aimed to characterize the pharmacokinetics and safety parameters of intravaginally-administered artesunate.

**Methods:** This is a Phase I, single-arm, open-label pharmacokinetic study of 12 healthy women in Kisumu, Kenya who self-administered 200 mg artesunate vaginal pessaries once daily for 5 consecutive days under direct observation with daily safety assessments. Participants returned each day for supervised dosing and adverse-effect assessments on days 1–4 without pharmacokinetic blood sampling. On day 5, participants administered the final dose and underwent serial blood sampling at 0.25, 0.5, 1, 2, 4, 6, and 8 hours. Plasma concentrations of artesunate and dihydroartemisinin (DHA), its active metabolite, were quantified and pharmacokinetic parameters estimated using non-compartmental analysis.

**Results:** Artesunate and its active metabolite dihydroartemisinin (DHA) were detectable in all participants, with mean (SD) C_max_ values of 83.7 (42.7) ng/mL and 97.0 (53.1) ng/mL, respectively, and a mean DHA AUC_(0-T)h_ of 504.2 (281.1) ng·h/mL. Additionally, the mean T_max_ values for Artesunate and DHA are 4.17(1.59) hours and 6.33 (1.67) hours respectively. Due to delayed absorption (T_max_), area under the serum concentration versus time curve (AUC), apparent clearance, and elimination half-life could not be calculated. The treatment was well tolerated with no serious adverse events.

**Conclusion:** Peak DHA concentrations (range 19.7–180.6 ng/mL) were substantially lower than those reported for intravenous and oral administration with interpatient variability (CV 54.8% for Cmax and 55.8% for AUC_0-T)_ hours) consistent with those reported for other non-intravenous routes of administration. Intravaginally-administered artesunate yielded low, yet measurable, systemic exposure with delayed T_max_. These findings support further investigation of intravaginal artesunate as a safe self-administered treatment for cervical precancer.

## Introduction

Cervical cancer is the fourth most common cancer among women worldwide and disproportionately burdens low- and middle-income countries (LMICs), where it remains the second leading cause of cancer-related mortality among women ^1^. In sub-Saharan Africa, the incidence and mortality from cervical cancer are among the highest globally, driven largely by limited access to vaccination, screening, and precancer treatment.^2^ While cervical cancer is preventable through HPV vaccination and detection and treatment of cervical precancer — high-grade cervical intraepithelial neoplasia (CIN2/3) — access to these interventions remains severely constrained in LMICs. Shortages of trained healthcare providers, inadequate health infrastructure, and the cost and distance required to reach referral centers leave many women unable to access timely care, underscoring an urgent need for more accessible and scalable, provider-independent cervical precancer treatment approaches.

Self-administered topical therapies represent a promising strategy to overcome these access barriers. Artesunate, a World Health Organization (WHO)-approved antimalarial agent derived from artemisinin^3^, has demonstrated anticancer activity through multiple mechanisms, including ferroptosis via its endoperoxide bridge reacting with intracellular ferrous iron, induction of cell cycle arrest, and modulation of pro-inflammatory and pro-proliferative pathways ^4–7^. HPV-infected epithelial cells overexpress the transferrin receptor and accumulate excess intracellular iron, rendering them potentially susceptible to artesunate-mediated cytotoxicity^4^. A multi-center, dose-escalation Phase I study in the United States demonstrated that intravaginal artesunate pessaries were safe, well-tolerated, and showed early efficacy signals for treatment of CIN2/3, with no grade 3 or 4 adverse events reported ^8^. These findings supported the advancement of artesunate to a randomized Phase II trial currently enrolling at multiple US sites (NCT04098744) and established the rationale for its evaluation in LMICs, where the drug is already widely available as an antimalarial.

A key unanswered question for deploying intravaginal artesunate in malaria-endemic settings is whether vaginal administration at the dose and frequency used for cervical precancer treatment results in meaningful systemic drug absorption. Artemisinin-based combination therapies (ACTs) are the WHO-recommended first-line treatment for uncomplicated *Plasmodium falciparum* malaria ^3^, and sub-therapeutic systemic exposure to artesunate could theoretically exert selective pressure on malaria parasites and contribute to artemisinin resistance — a growing public health concern in Africa^9,10^. While the pharmacokinetics (PK) of artesunate and its active metabolite dihydroartemisinin (DHA) have been well characterized following intravenous, intramuscular, oral, and rectal administration ^11–13^, no data exist on systemic absorption following intravaginal use. The cervicovaginal epithelium, a thick stratified squamous epithelium that is significantly less vascular than the rectal mucosa, may substantially limit systemic uptake of intravaginal artesunate.

To address this evidence gap, we conducted a Phase I pharmacokinetic study of intravaginal artesunate in healthy women in Kisumu, Kenya. The primary objective was to characterize systemic exposure to DHA — quantified as area under the serum concentration-time curve (AUC) — following five consecutive days of self-administered 200 mg artesunate vaginal pessaries at the dose and frequency intended for cervical precancer treatment. Secondary objectives included assessment of additional PK parameters for artesunate and DHA, as well as evaluation of safety.

## Methods

This was a single-arm, open-label Phase I pharmacokinetic study, conducted in Kisumu, Kenya, at Lumumba Sub-County Hospital. The primary objective was to characterize the serum pharmacokinetics (PK) of artesunate and its dihydroartemisinin (DHA) metabolite following five consecutive days of self-administered intravaginal artesunate. The recruitment period started on May 29, 2024, and ended on August 1, 2024.

Twelve healthy female participants aged 18 to 65 years were enrolled and instructed to self-administer a 200 mg artesunate vaginal pessary once daily for five consecutive days under direct observation at the study clinic. Eligibility criteria included a body weight of ≥50 kg, negative malaria antigen and pregnancy tests, willingness to use hormonal or barrier contraception during the 5-day dosing period (for participants of childbearing age, defined as under 50 years), and plans to reside locally throughout the study duration. Participants were excluded if they were pregnant or breastfeeding, currently using or had recently used (within 3 days) artemisinin-based therapy, taking efavirenz, assigned male at birth, had a history of total hysterectomy, tested positive for malaria at screening, or had any comorbid condition that could interfere with study participation, as determined by the investigator.

Participants underwent pre-screening and provided written informed consent prior to enrollment. Screening procedures included a review of medical history, pelvic examination, urine pregnancy test, malaria antigen test, and instruction on self-administration of artesunate using a pelvic model. On Visit 1 (day 1), a baseline blood sample (2.5 mL) was collected before the first self-administered 200 mg artesunate vaginal pessary. Visits 2–4 (days 2–4) involved daily return for supervised dosing and assessment of adverse events using the Common Terminology Criteria for Adverse Events (CTCAE). On Visit 5 (day 5), a pre-dose blood sample at time 0 was collected followed by the final dose of artesunate; serial blood samples were then obtained at 15 minutes, 30 minutes, 1, 2, 4, 6, and 8 hours post-dose for PK analysis. Visit 6 occurred between days 9–15 for follow-up safety assessment and documentation of any delayed adverse events. Further study details are available in the published protocol.^14^

PK parameters were calculated using non-compartmental analysis of Day 5 serum concentration-time data. The following PK parameters were estimated for artesunate and DHA maximum concentration (C_max_, time to C_max_ (T_max_)), last concentration (C_last_), time of C_last_ (T_last_)) and area under the concentration versus time curve from 0 to T_last_ (AUC_0-t_). Due to a lack of an elimination phase in the concentration versus time profile, the following PK parameters could not be estimated apparent clearance (CL/F), apparent volume of distribution (Vd/F), and half-life (t□/□).

The primary endpoint, mean DHA AUC_0-t_, was compared to historical PK data from intravenous, oral, and rectal routes using a one-sample Student’s t-test (p<0.05). Other PK parameters were summarized descriptively and interpreted in the context of existing literature. Safety was monitored throughout the study, and all adverse events (AEs) were graded according to CTCAE v5.0.

### Ethical Considerations

The study protocol was approved by the University of North Carolina-Chapel Hill Institutional Review Board and the African Medical Research Foundation Ethics Review Committee. All participants provided written informed consent prior to enrollment. The trial is registered at ClinicalTrials.gov (NCT06263582).

## Results

### Participant characteristics

Participant characteristics are described in **Table 1**. Participants in this study had completed at least secondary education, and most were current students. Their mean age was 23.3 (SD 5.1), on average they had each had one pregnancy, and none were married.

**Table 1.** Demographic characteristics of participants in a pharmacokinetic study of intravaginal artesunate in Kisumu, Kenya.

| Characteristics | Enrolled Participants n = 12 |
| --- | --- |
|  | Mean (SD) |
| Age, years | 23.3 (5.1) |
| Gravidity | 1 (0) |
| Parity (Live births) | 1 (0) |
| **Level of education** | n (%) |
| Completed Secondary | 10 (83.3) |
| Completed College | 2 (16.7) |
| **Occupation** |  |
| Salaried | 2 (16.7) |
| Student | 10 (83.3) |
| **Daily Income** |  |
| <Ksh500 | 10 (83.3) |
| >Ksh500 | 2 (16.7) |
| **Marital Status** |  |
| Single/Never Married | 12 (100.0) |
| **Electricity available** |  |
| Yes | 11 (91.7) |
| No | 1 (8.3) |
| **Tap water available** |  |
| Yes | 10 (83.3) |
| No | 2 (16.7) |
| **Current or prior tobacco use** |  |
| Yes | 2 (16.7) |
| No | 10 (83.3) |
| **Current or prior alcohol use** |  |
| Yes | 9 (75.0) |
| No | 3 (25.0) |
| **Current contraception use** |  |
| Yes | 12 (100.0) |
| **Method of contraception** |  |
| Intrauterine contraceptive device | 1 (8.3) |
| Condoms | 11 (91.7) |

### Pharmacokinetics

Following intravaginal administration of a single 200 mg dose of artesunate on day 5, plasma pharmacokinetics were characterized for artesunate and its active metabolite dihydroartemisinin (DHA) over an 8-hour period in 12 participants. Systemic absorption of artesunate and subsequent metabolic conversion to DHA were observed in all individuals with interindividual variability in exposure levels.

The concentration versus time profiles for artesunate are shown in **Fig 1**, with corresponding pharmacokinetic parameters summarized in **Table 2**. Artesunate appeared in plasma rapidly, with quantifiable concentrations observed in most participants within 15–30 minutes post-dose. The mean (SD) C_max_ was 83.7 (42.7) ng/mL, representing a 6.3-fold difference across participants. C_max_ of artesunate was reached at a mean (SD) T_max_ of 4.17 (1.59) hours (range 2-8 hours). Artesunate concentrations plateaued in most individuals and measurable levels persisted through the final sample time point of 8 hours post-dose. The mean (SD) artesunate concentration at 8 hours (C_last_); was 62.7 (34.50) ng/mL (range 14.45-115.82). Systemic exposure over time was assessed using the area under the plasma concentration versus time curve from 0 to 8 hours (AUC_0-t_). The mean (SD) artesunate AUC_0-t_ was 464.9 (228.7) ng·h/mL (range 145.87-824.15).

**Table 2.**
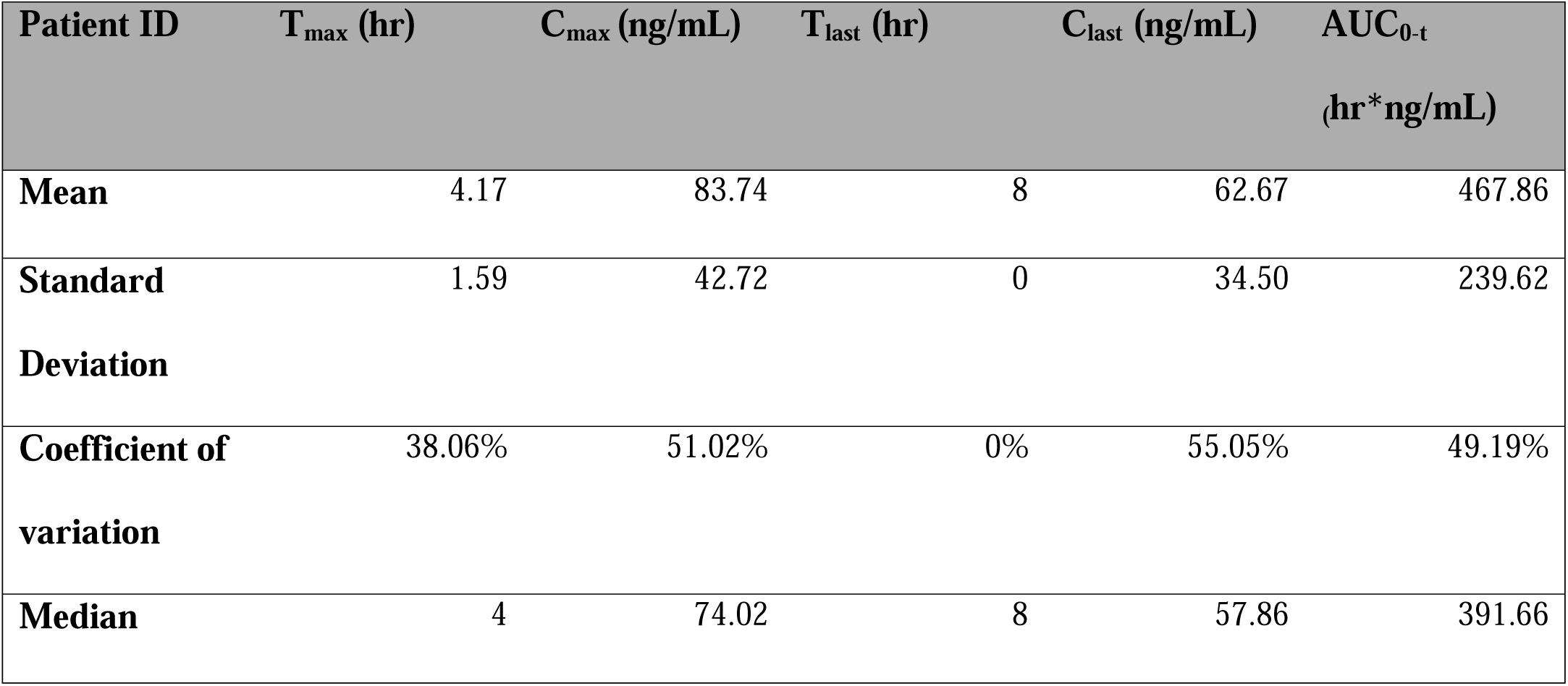
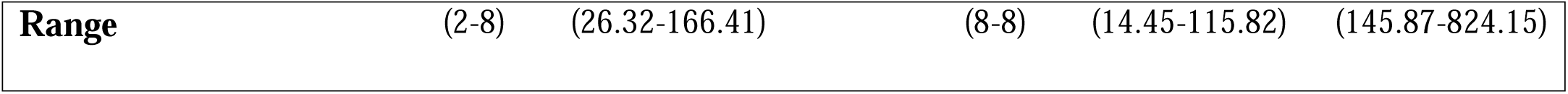
Summary of Artesunate Plasma Pharmacokinetic Parameters. *T_max_,* time to maximum concentration; *C_max_,* maximum concentration; *T_last_*time of last measurable concentration; *C_last_*, last measurable concentration; *AUC*, area under curve.

The concentration versus time profiles for DHA are shown in **Fig 2**, with corresponding pharmacokinetic parameters summarized in **Table 3**. DHA was detected in plasma following a lag period, consistent with its formation via metabolic conversion from artesunate. Peak plasma concentrations (C_max_) of DHA were reached at a mean (SD) T_max_ of 6.33(1.67) hours (range 4–8 hours). The mean (SD) C_max_ was 97.0 (53.1) ng/mL (range of 19.7–180.6 ng/mL), representing a 9.2-fold difference across participants. DHA concentrations continued to rise or remained relatively stable through 8 hours sampling schedule in most individuals. The mean (SD) of DHA Clast at 8 hours was 85.6 (52.8) ng/mL (range 19.7–180.6 ng/mL). The mean (SD) for DHA AUC_(0-T) h_ was 504.2 (281.1 ng·h/mL) and the values ranged from 108.5 to 912.3 ng·h/mL). The mean (SD) of the ratio of DHA AUC_0-t_ to artesunate AUC_0-t_ was 1.08 (0.27) and values ranged from 0.50–1.55. These findings support DHA’s prolonged systemic availability after intravaginal administration of artesunate.

**Table 3.** Summary of active metabolite dihydroartemisinin (DHA) parameters in plasma T_max_, time to maximum concentration; C_max_, maximum concentration; T _last,_ last time of last measurable concentration; C_last_, last measurable concentration; AUC, area under curve.

| Patient ID | T<sub>max</sub> (hr) | C<sub>max</sub> (ng/mL) | T<sub>last</sub> (hr) | C<sub>last</sub> (ng/mL) | AUC<sub>0-t</sub><br>(hr*ng/mL) |
| --- | --- | --- | --- | --- | --- |
| **Mean** | 6.33 | 97.04 | 8 | 89.93 | 504.21 |
| **Standard Deviation** | 1.67 | 53.12 | 0 | 53.12 | 281.11 |
| **Coefficient of Variation** | 0.26 | 0.55 | 0 | 0.59 | 0.557526 |
| **Median** | 6 | 90.18 | 8 | 81.81 | 420.1 |
| **Range** | (4,8) | (19.68,180.63) | 8 | (19.68,180.63) | (108.49, 912.27) |

No clear associations were observed between participant characteristics and exposure.

Following intravaginal administration of artesunate, mean C_max_ (SD) values were 83.7 (42.7) ng/mL for artesunate and 97.0 (53.1) ng/mL for DHA. These values are substantially lower than those observed with intravenous artesunate at 2 mg/kg (artesunate: 19,420 ng/mL; DHA: 1,286 ng/mL) and 4 mg/kg (artesunate: 36,100 ng/mL; DHA: 3,148 ng/mL). Compared with other non-intravenous routes, exposure also remained lower than oral administration at a dose of 200 mg artesunate (artesunate: 256.3 (167.8) ng/mL; DHA: 873.7 (366) ng/mL) and rectal administration at a dose of 200 mg artesunate (artesunate: 448.5 (286.7) ng/mL; DHA: 385.6 (173.9) ng/mL).^15–18^ Mean AUC_(0-T)h_ values were also lower than those reported for intravenous, oral, and rectal administration^17^, where rectal artesunate (200 mg) produced an AUC of 796 (423) ng·hr/mL for artesunate and an AUC – of 965 (607) ng·hr/mL for DHA, as demonstrated in **Table 4**.

**Table 4.** One-sample one-tailed Student’s t-test comparing observed intravaginal DHA AUC against mean DHA AUCs by route of administration.

| Route of administration | Hypothesized mean<br>(ng·hr/mL) | t-statistic | df | p-value (one-tailed) |
| --- | --- | --- | --- | --- |
| Intravenous (IV) 4<br>mg/kg | 4886.0 | -54.00 | 11 | 1.000 |
| Rectal 200 mg | 965 | -5.678 | 11 | 1.000 |
| Oral 200 mg | 1331 | -10.188 | 11 | 1.000 |

Observed sample: mean DHA AUC = 504.21 ng·hr/mL, SD = 281.11, n = 12 (df = 11, SE = 81.15 ng·hr/mL). H<sub>0</sub>: observed mean > hypothesized mean. Statistical significance set at $\alpha = 0.05$ . No comparisons reached statistical significance. DHA, dihydroartemisinin; AUC, area under the concentration–time curve; IV, intravenous; df, degrees of freedom.

### Safety

A total of 13 AEs were reported in this study, which are summarized in **Table 5**. The majority of AEs (n=11, 84%) were Grade 1. The commonly reported AEs included mild abdominal pain (n=3), abnormal vaginal discharge (n=6), and vaginal itching (n=2), all assessed as related to the investigational product. A Grade 2 vaginal lesion reported by a participant was considered possibly related to treatment. No serious AEs occurred, and all participants completed the study without discontinuation due to AEs.

**Table 5.** Type, frequency, and duration of adverse events (AEs) among participants of a Phase 1 pharmacokinetic trial of intravaginal artesunate.

| Reported Adverse Event | Grade 1 (n %) | Grade 2 (n %) | Mean Duration (days), SD |
| --- | --- | --- | --- |
| Abdominal pain | 3(25.0%) | - | 1, 0 |
| Abnormal vaginal discharge | 6(50.0%) | - | 1,0 |
| Pain on the left upper arm<sup>1</sup> | - | 1(8.3%) | 6,0 |
| Vaginal lesions<sup>2</sup> |  | 1(8.3%) | 5,0 |
| Vaginal itching | 2(16.7%) | - | 2,1.4 |

1. Pain in the left upper arm was accompanied by an elbow wound seen on examination. This adverse event was unrelated to the study product.
2. Vaginal lesions were confirmed on pelvic exam.

## Discussion

To our knowledge, this is the first study to characterize plasma pharmacokinetics following intravaginal artesunate administration. The study also demonstrated that intravaginal artesunate was safe and well-tolerated, as all adverse events were Grade 1 or Grade 2 in severity with no severe adverse events reported. Artesunate and DHA were detectable in serum in all participants after intravaginal administration. Artesunate was detectable in plasma within 15 to 30 minutes post-dose, similar to reported T_max_ values of 0.58−1.43 hours following rectal administration of artesunate.^16^ DHA appeared after a lag period consistent with metabolic conversion from artesunate through esterase-catalyzed hydrolysis in the bloodstream.^16^ These lower absorption rates likely occur because the vaginal mucosa relies on passive diffusion through multiple cell layers and lacks the active transport systems or villi found in highly absorptive tissues.^19^ Therefore, both the rate and extent of systemic absorption are lower than those achieved with oral administration or direct intravenous injection.

There was substantial interpatient variability in plasma exposure of artesunate and DHA, with coefficients of variation exceeding 49–55% for C_max_ and AUC _(0-t)_ hours. When contextualized against other routes, similar or greater variability has been reported in primary pharmacokinetic studies of artesunate and dihydroartemisinin pharmacokinetics across non-intravenous route**s** of administration. For example, oral artesunate demonstrated a C_max_ of 256.3 (167.8) ng/mL for artesunate and 873.7 (366) ng/mL for DHA,^17^ corresponding to coefficients of variation of approximately 65.5% and 41.9%, respectively. Rectal administration of artesunate similarly showed marked dispersion, with coefficients of variation approximating 63.9% and 45.1%.^17^ Intramuscular administration of artesunate in participants with uncomplicated *Plasmodium falciparum* malaria also demonstrated wide concentration ranges.^18^ Collectively, these findings indicate that coefficients of variation in the 50% range are not abnormally high for artesunate and DHA across other non-intravenous routes but rather fall within the expected moderate-to-high interpatient variability observed in prior clinical pharmacokinetic studies.

However, variability estimates across studies may not be directly comparable due to differences in study populations, dosing regimens, and reporting methods. The variability in plasma exposure of artesunate and DHA for vaginally administered artesunate may result from individual differences in vaginal mucosal permeability, hormonal status, age-related epithelial changes, enzymatic conversion efficiency, and systemic clearance.^20,21^

Concentration–time profiles demonstrate delayed, and sustained absorption of artesunate and DHA following intravaginal administration compared with intravenous, rectal, and oral routes, as evidenced by a mean T_max_ of Artesunate of 4.17 hours in the present study versus 1.43 and 0.66 hours following rectal and oral administration, respectively.^15,17^ As mean T_max_ of DHA was 6.33(1.67) hours, conversion from artesunate to DHA took from 0 to 2 hours, with prolonged detectable concentrations from 0 to 8 hours and no clearly defined elimination phase within the sampling window, thus limiting potential comparisons with other routes of administration. Additionally, C_last_ of artesunate and DHA were 62.67 (34.4) ng/mL and 85.62 (52.8) ng/mL, respectively, demonstrating prolonged detectable concentrations within the 8-hour sampling window, likely also related to the slow, but sustained absorption through vaginal mucosa.

These data raise the important question of whether systemic exposure following intravaginal dosing of artesunate could contribute to artemisinin resistance. Resistance is characterized clinically by delayed parasite clearance (following treatment with artesunate monotherapy or Artemisinin-based combination therapies).^9^ The primary drivers of resistance, both *in vitro* and *in vivo,* are point mutations in the *P. faclciparum* Kelch13 (*PfK13)* such as C580Y, A675V, R561H, and C469Y.^9,22–24^ Laboratory studies using the ring-stage survival assay (RSA) expose parasites to DHA concentrations of approximately 700 nM (∼200 ng/mL) for 4–6 hours, a dose that represents high, clinically relevant drug exposure that makes it easier to distinguish resistance parasites by their survival following short-term treatment.^25^ Resistant strains exhibit survival rates >1%, whereas sensitive strains show near-complete clearance.^25^ Importantly, the RSA models the precise pharmacologic scenario in which transient, subtherapeutic exposures may provide the selective pressure that enables resistant parasites to persist and propagate.

In this study, the mean DHA Cmax was 97 ng/mL, with a maximum individual value of 181 ng/mL. These concentrations approach, but do not exceed, the ∼200 ng/mL threshold used in RSA assays to define reduced susceptibility.^25^ Although overall exposure was markedly lower than therapeutic antimalarial dosing, the proximity of some individual concentrations to the RSA benchmark warrants caution, especially if dosing regimens increase cumulative or sustained exposure to artesunate, to ensure that systemic exposure does not approach or exceed thresholds associated with parasite survival. However, the much lower AUC of DHA compared to oral, rectal, and IV routes of administration is reassuring.

A key limitation of this study is that the sampling window was restricted to 8 hours, given that the known half-life of oral and rectal artesunate is 0.5–1.5 and 0.9–0.95 hours respectively.^16^ However, this precluded reliable estimation of terminal elimination parameters, including half-life, clearance, volume of distribution, and AUC –∞, as the Tmax of intravaginal artesunate was delayed.

Though low systemic absorption is demonstrated within 8 hours, future PK studies of artesunate following intravaginal administration should extend sampling to 24–48 hours to fully characterize elimination and total systemic exposure. Future studies should also consider the impact of vaginal dysbiosis on systemic absorption of intravaginal medications, especially in sub-Saharan Africa, where the prevalence of bacterial vaginosis is 20–50%.^26^ Trials ongoing for efficacy and feasibility of intravaginal artesunate should closely monitor malaria incidence within the study cohort. If found effective, self-administered topical therapies can be transformative in increasing access to secondary prevention of cervical cancer for marginalized women globally.

## Data Availability

The datasets generated and/or analyzed during the current study are available from the corresponding author upon reasonable request.

## Acknowledgements

The authors are grateful to the participants of the phase 1 study of intravaginal artesunate for their valuable contributions to our understanding of this medication in the context of their lives. We also thank the research teams at KEMRI for their support at the study site. Artesunate vaginal suppositories (pessaries) were provided as in-kind support by Frantz Viral Therapeutics (Mentor, OH).

---

**Fig 1.** The plasma concentration versus time profile of artesunate for each patient on day 5 of artesunate administration. The results for each patient are represented by a separate same color with measured concentrations depicted as the symbol and connected by lines.

**Figure 2.** The plasma concentration versus time profile of DHA for each patient on day 5 of artesunate administration. The results for each patient are represented by a separate same color with measured concentrations depicted as the symbol and connected by lines.

